# A comparative analysis of pediatric mental health-related emergency department utilization in Montréal, Canada before and during the COVID-19 pandemic

**DOI:** 10.1101/2022.04.18.22273970

**Authors:** Gabrielle Beaudry, Olivier Drouin, Jocelyn Gravel, Anna Smyrnova, Andreas Bender, Massimiliano Orri, Marie-Claude Geoffroy, Nicholas Chadi

**Affiliations:** Department of Psychiatry, University of Oxford, Oxford (UK); Sainte-Justine Hospital Research Center, Montréal, Québec (Canada); Department of Pediatrics, Université de Montréal, Montréal, Québec (Canada); Division of General Pediatrics, Sainte-Justine University Hospital Centre, Montréal, Québec (Canada); Department of Social and Preventive Medicine, School of Public Health, Université de Montréal, Montréal, Québec (Canada); Division of Pediatric Emergency Medicine, Sainte-Justine University Hospital Centre, Montréal, Québec (Canada); Department of Statistics, LMU Munich, Munich (Germany); McGill Group for Suicide Studies, Douglas Mental Health University Institute, Department of Psychiatry, McGill University, Montréal, Québec (Canada); Department of Educational and Counselling Psychology, McGill University, Montréal, Québec (Canada); Division of Adolescent Medicine, Sainte-Justine University Hospital Centre, Montréal, Québec (Canada)

## Abstract

**Background:** Reports on longitudinal trends in mental health–related (MHR) emergency department (ED) utilization spanning the pre- and post-pandemic periods are lacking, along with evidence comparing healthcare services utilization by sociodemographic subgroups. The aim of this study was to evaluate COVID-19–associated changes in MHR ED utilization among youth overall and by age, sex, and socioeconomic status (SES).

**Methods:** This retrospective cross-sectional study analyzed MHR ED utilization before and during the COVID-19 pandemic at a large urban pediatric tertiary care hospital in Montréal, Canada. All ED visits for children (5–11 years) and adolescents (12–17 years) between April 1, 2016 and November 30, 2021 were included. The main outcome was the monthly count of MHR ED visits. Pre-pandemic and pandemic periods were compared using an interrupted time series design. The effect of seasonality (in months), age (in years), sex (male or female), and SES (low, average, high) were compared using a generalized additive model.

**Results:** There were a total of 437,147 ED visits (204,215 unique patients) during the five-year study period of which 9,748 (5.8%) were MHR visits (7,686 unique patients). We observed an increase of 69% (95% CI, +53% to +85%; p = .001) in the mean monthly count of MHR ED visits during the pandemic period, which remained significant after adjusting for seasonality (44% increase, 95% CI, +38% to +51%; p = .001). The chance of presenting for a MHR ED visit increased non-linearly with age. There were increased odds of presenting for a MHR ED visit among girls between the pre-pandemic and pandemic periods (OR 1.42, 95% CI 1.29–1.56). No difference by SES group during and before the COVID-19 pandemic was found (OR 1.01, 95% CI 0.89–1.15 [low]; OR 1.09, 95% CI 0.96–1.25 [high]).

**Conclusions:** Our study shows important increases in MHR ED utilization among youth, and especially among girls, during the first 20 months of the COVID-19 pandemic, highlighting the need for sustained, targeted and scalable mental health resources to support youth mental health during the current and future crises.

## BACKGROUND

Since COVID-19 was declared a global pandemic,(1) public health measures implemented to reduce transmission of the virus have led to considerable changes in the delivery of pediatric healthcare services.(2) Delivery modes, such as telemedicine, rapidly gained importance,(3, 4) while utilization of most in-person services plummeted in the first months of the pandemic.(5) Large fluctuations in pediatric emergency department (ED) utilization have been observed.(6-17) Pediatric EDs are integral to the assessment, treatment and coordination of care for children and adolescents, and often serve as a safety net for vulnerable and underserved patients.(18) Increasingly, EDs also play a pivotal role in pediatric mental health emergencies, as exemplified by the rising pre-pandemic trends in mental health-related (MHR) ED visits documented in the last decade.(19, 20)

Reported number and proportion of MHR ED visits have not consistently followed the pattern of overall pediatric ED visits following the pandemic onset. While both MHR and overall ED visits first decreased during 2020,(7-12, 14-17, 21-24) some studies have reported increased MHR ED utilization during 2021 and early 2022.(13, 25) During this time, overall pediatric ED utilization has remained lower than pre-COVID-19 levels.(6, 25)

So far, the bulk of the evidence regarding changes in pediatric MHR ED utilization has originated from the United States, thereby limiting the generalisability of findings to other countries. Potential regional and national differences could stem from varying COVID-19 epidemiology, public health responses, healthcare systems, socioeconomic and sociocultural factors, and availability of mental health services.(26) Moreover, it remains unclear whether MHR ED utilization has differed across pediatric subpopulations. To date, only two studies have examined sociodemographic-specific differences — notably, related to age, sex and socio-economic status (SES).(13, 25) The first was conducted in two large pediatric centers in New South Wales, Australia, and found increased MHR ED utilization among youth between June 2020 and February 2021, with higher increases among girls and children from socioeconomically advantaged areas.(13) The second study, which analyzed data from the National Syndromic Surveillance Program in the US, showed that adolescent girls aged 12–17 years accounted for the largest increases in both the number and proportion of MHR ED visits seen in 2020, 2021 and January 2022 when compared to 2019.(25) Additional research is required to prevent further exacerbation of intersectional inequalities in youth mental health,(27) and to inform effective service planning and resource allocation in pediatric emergency care.

In this study, we aimed to identify changes in MHR ED utilization before and during the COVID-19 pandemic in Montréal, Canada. We further sought to determine whether COVID-19 showed differential associations by sociodemographic group, with respect to age, sex, and SES.

## METHODS

### Study design, setting and population

For this retrospective cross-sectional study, we used an interrupted time series (ITS) design to compare MHR ED utilization before and during COVID-19 at the *Centre hospitalier universitaire* (CHU) Sainte-Justine. The CHU Sainte-Justine is a high-volume tertiary pediatric university hospital located in Montréal (Québec), Canada. The study population was comprised of all ED visits for children (5–11 years) and adolescents (12–17 years) between April 1, 2016, and November 30, 2021. We excluded ED visits for younger children (<5 years) as psychiatric consultation or treatment is uncommon in this age group.(19) The primary cohort was defined as all MHR ED visits with complete patient information. Visits for which patients left the hospital prior to receiving care were excluded due to lack of diagnosis. Ethical approval was obtained from the CHU Sainte-Justine Research Ethics Committee (Protocol ID: MP-21-2021-2930). All patient data were deidentified; thus, informed consent was not required. We followed the Strengthening the Reporting of Observational Studies in Epidemiology (STROBE) reporting guideline.(28)

### Outcomes and variables

#### Exposure

The main exposure was a dummy variable coding the pre-pandemic (0) and the pandemic period (1). We defined the start of the pandemic period as March 1, 2020, consistent with the initial implementation of COVID-19–related public health measures in the province of Québec.(29) Thus, the pre-pandemic period spanned from April 1, 2016, to February 29, 2020, whereas the pandemic period spanned from March 1, 2020 to November 30, 2021.

#### Outcomes

The outcomes of the ITS analysis included the monthly count of MHR ED visits stratified into the pre-pandemic and pandemic periods. The outcome for the generalized additive model (GAM) was the type of ED visits, being MHR visits or non-MHR visits.

#### Sociodemographic characteristics

We employed a computer query tool (B-Care) to interrogate various hospital administrative databases. We focused on a patient-level ED database that includes demographic (sex, age and 6-digit postal code) and clinical (primary discharge diagnosis) information. The primary discharge diagnosis — recorded by the treating physicians based on a local list of 658 diagnoses — was used to determine the type of ED visits. We report the list of included MHR ED diagnoses in eTable 1.

**Table 1.** Characteristics of ED visits, April 1, 2016 to November 2021.

|  | Prepandemic | Pandemic |  |  |  |
| --- | --- | --- | --- | --- | --- |
| All ED visits | 2016-2019<br>(N=118728) | 2020<br>(N=23317) | 2021<br>(N=26704) | Overall<br>(N=168749) |  |
| MHR visits | 5927 (5.0%) | 1521 (6.5%) | 2300 (8.6%) | 9748 (5.8%) |  |
| Other visits | 112801 (95.0%) | 21796 (93.5%) | 24404 (91.4%) | 159001 (94.2%) |  |
| MHR ED visits | 2016-2019<br>(N=5927) | 2020<br>(N=1521) | 2021<br>(N=2300) | Overall<br>(N=9748) | p-value |
| **Age (continuous)** |  |  |  |  |  |
| Median (IQR) | 14.3 (11.0-16.0) | 14.4 (11.7-16.1) | 14.4 (12.2-15.9) | 14.3 (11.4-16.0) | 0.045 |
| **Age group** |  |  |  |  |  |
| 5–11 years | 1803 (30.4%) | 413 (27.2%) | 547 (23.8%) | 2763 (28.3%) | <0.001 |
| 12+ years | 4124 (69.6%) | 1108 (72.8%) | 1753 (76.2%) | 6985 (71.7%) |  |
| **Sex** |  |  |  |  |  |
| Female | 3521 (59.4%) | 970 (63.8%) | 1615 (70.2%) | 6106 (62.6%) | <0.001 |
| Male | 2406 (40.6%) | 551 (36.2%) | 685 (29.8%) | 3642 (37.4%) |  |
| **SES** |  |  |  |  |  |
| Most privileged | 1918 (32.4%) | 514 (33.8%) | 805 (35.0%) | 3237 (33.2%) | 0.087 |
| Average | 1009 (17.0%) | 252 (16.6%) | 408 (17.7%) | 1669 (17.1%) |  |
| Most deprived | 2755 (46.5%) | 675 (44.4%) | 972 (42.3%) | 4402 (45.2%) |  |
| Missing | 245 (4.1%) | 80 (5.3%) | 115 (5.0%) | 440 (4.5%) |  |

Note. Data are n (%). Counts include multiple visits for the same patient. Between-time period differences were assessed using Kruskal-Wallis (for continuous variables) and $\chi^2$ tests (for categorical variables).
Abbreviations: ED, emergency department; MHR, mental health-related; SES, socioeconomic status.

Patients’ sex assigned at birth was coded as male or female. Patients’ age at diagnosis was considered both as a continuous and as a dichotomous variable (5–11 years vs 12–17 years). We selected an age cut-off at 12 years to reflect the difference between primary (elementary school) and secondary (middle and high school) education, as public health measures have differed across levels of education in Québec. SES was determined by linking the patients’ postal code at the time of diagnosis with Statistics Canada’s Postal Code Conversion File, thus allowing us to identify the relevant dissemination area based on the most recent Canadian Census (from 2016). We then linked this area-level information to Pampalon’s material deprivation index, which combines measures of income, employment, and education. According to this index, SES is categorized into quintiles from Q1 (least materially deprived) to Q5 (most materially deprived).(30) Based on previous literature, we combined the two lowest and the two highest quintiles to create three distinct material deprivation profiles: most privileged (Q1–Q2), average (Q3), and most deprived (Q4–Q5).(31)

### Statistical analysis

We first used descriptive statistics to summarize patient characteristics for MHR ED visits. Kruskal-Wallis and χ^2^ tests were employed to compare between-time period differences for continuous and categorical variables, respectively. To investigate changes in MHR ED utilization between the pre-pandemic and pandemic periods, we conducted an ITS analysis of aggregated monthly MHR ED visits using a Bayesian structural time series model (BSTS). We treated the monthly count of MHR ED visits as a time series and the month at which the exposure first occurred as the event of interest. The outcome was decomposed into trend-cycle, seasonal, and remainder components. The seasonal component was then removed to obtain a time series adjusted for seasonal variations in MHR ED visits.

To investigate associations between sociodemographic characteristics and the outcome, a generalized additive model (GAM) with binomial response and a logistic link was specified. GAMs are an extension of generalized linear models (GLMs), whereby predictors are linked to the outcome using smooth functions, allowing for greater flexibility in incorporating nonlinear forms of the predictors.(32) Model parameters were estimated via restricted maximum likelihood (REML).(33) The following covariates were included in the model: seasonality (in months) and age (in years), continuous variables modeled as semi-parametric smoothed terms; COVID-19 exposure (the main exposure), sex, and SES, categorical variables modeled as parametric linear terms. For the smoothed terms, we specified factor-smooth interactions, by which a separate smooth function was estimated for each level of COVID-19 exposure (i.e., pre-pandemic and pandemic periods). An interaction between linear terms and COVID-19 exposure was also specified. As part of a sensitivity analysis, we fitted another model with age as a dichotomous variable.

Analyses were performed using R version 4.1.2 (R Project for Statistical Computing). ITS analysis was conducted using the CausalImpact and seasonal packages.(34, 35) We used the mcgv package for GAM estimation.(33) Statistical significance was set at two-sided p-value <.05.

## RESULTS

Between April 2016 and November 2021, there were a total of 437,147 ED visits (204,215 unique patients). Among these visits, children and adolescents aged 5 to 17 years accounted for 168,749 ED visits (99,313 unique patients), of which 9,748 (5.8%) were MHR visits (7,686 unique patients). Patient characteristics are summarised in Table 1. We found significant differences between time periods (pre-pandemic [2016–2019] and pandemic [2020, 2021]) for all patient characteristics, with the exception of SES. There were missing data for the SES (approximately 5% of visits per year), and the age of patients in the included MHR ED visits was significantly higher than that of the excluded ones (eTable 2). The monthly count of MHR ED visits is presented by time period and sociodemographic characteristics in Figures 1 and 2, respectively. The frequency of MHR ED visits per unique patient can be found in the Supplement (eTable 3).

**Figure 1.**
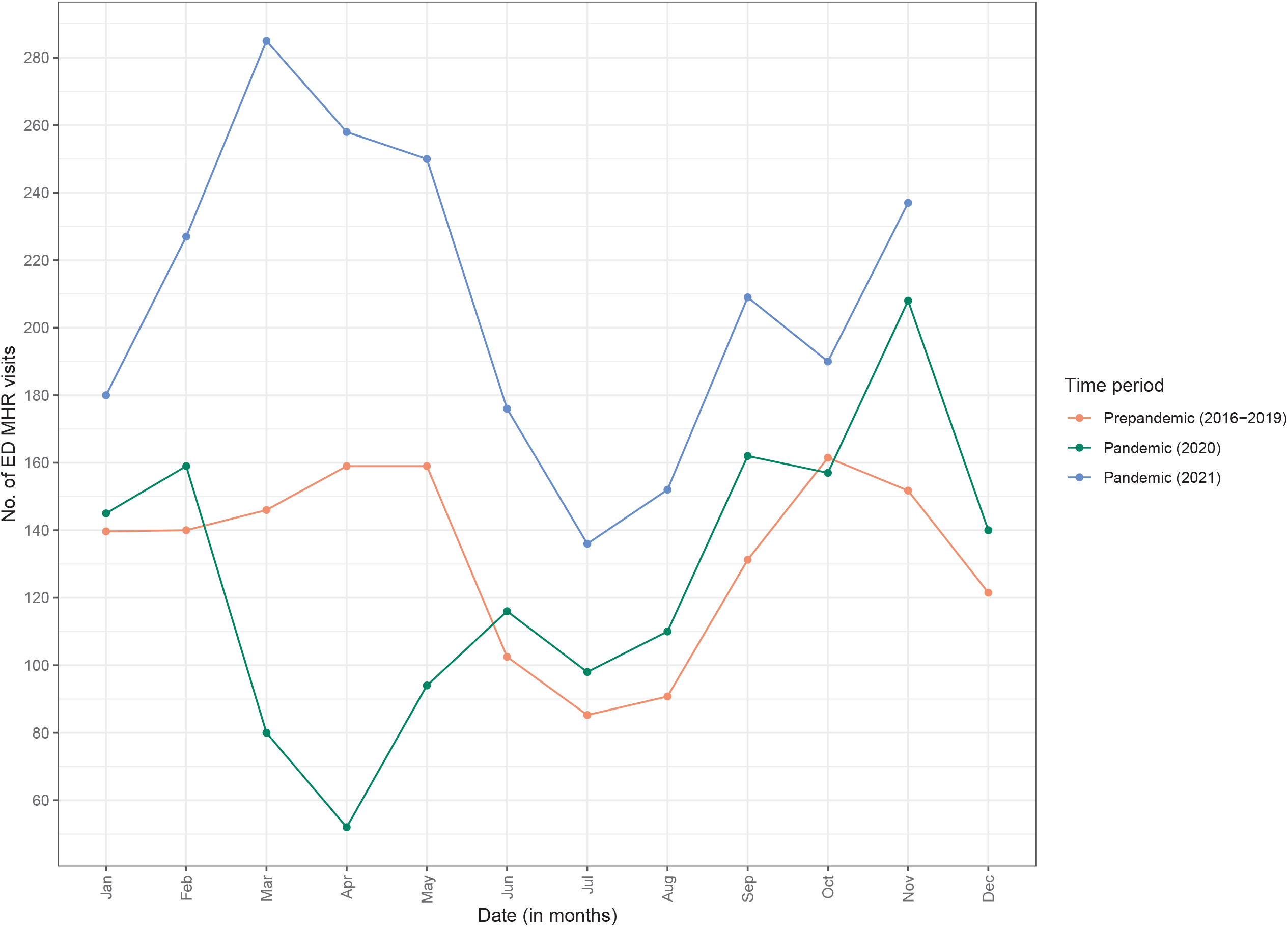
MHR ED utilization at the CHU Sainte-Justine from April 1, 2016 to November 30, 2021 (by time period) Abbreviations: CHU, *centre hospitalier universitaire*; ED, emergency department; MHR, mental health-related.

**Figure 2.**
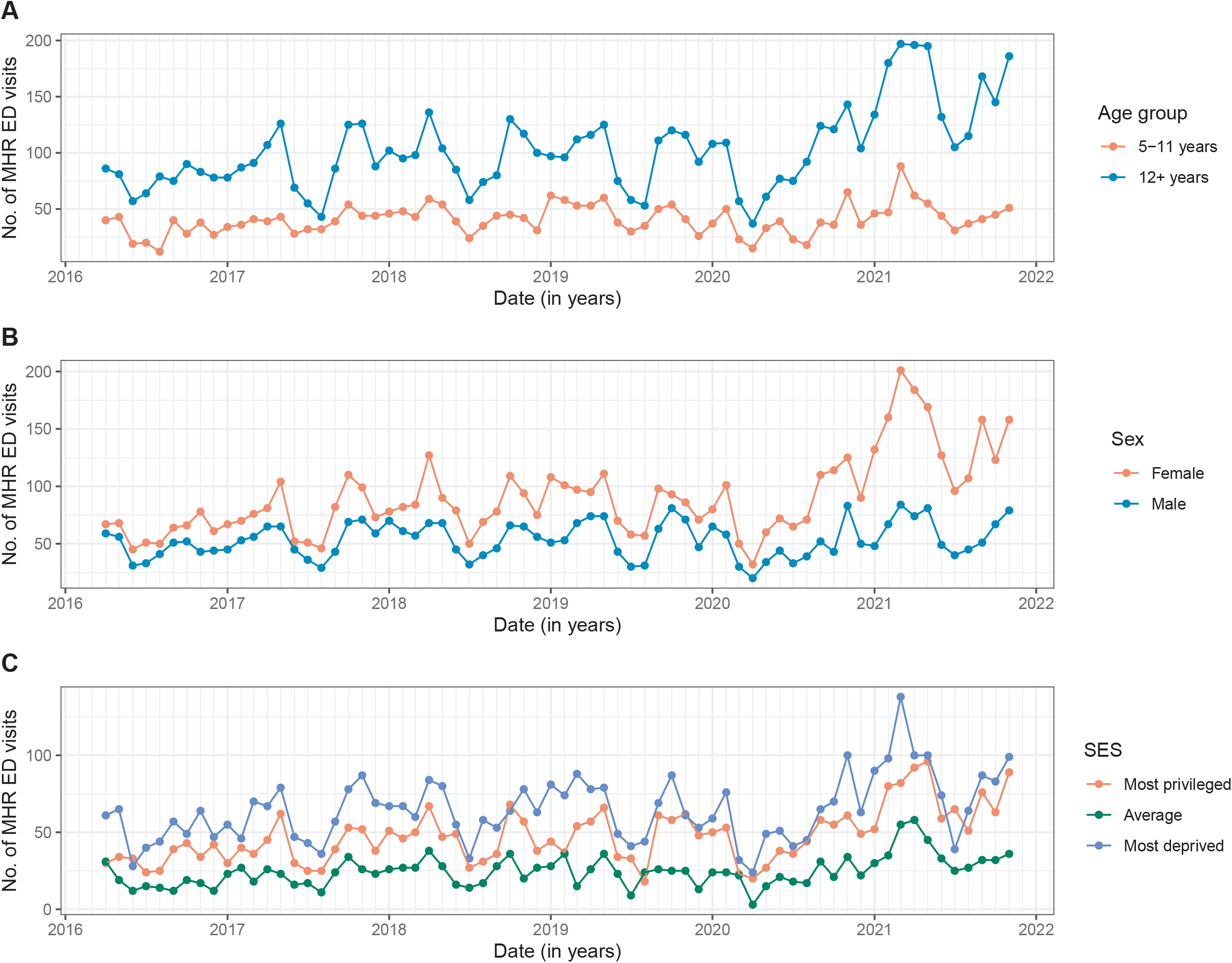
MHR ED utilization at the CHU Sainte-Justine from April 1, 2016 to November 30, 2021 (by age group, sex and SES) Note. Panels A, B and C show the time series of the monthly number of MHR ED visits by age group, sex and SES, respectively. Abbreviations: CHU, *centre hospitalier universitaire*; ED, emergency department; MHR, mental health-related; SES, socio-economic status.

Bayesian structural time series analysis revealed that after an initial decrease in MHR ED visits between March and June 2020, monthly visit counts exceeded those from pre-pandemic years as shown in Figure 3 (deseasonalized data), eFigures 1-2 (raw data) and eFigure 3 (raw and deseasonalized data). Overall, we observed an increase of 69% (95% CI, +53% to +85%; *p* = .001) in the mean monthly count of MHR ED visits, which remained significant after adjusting for seasonality (44% increase, 95% CI, +38% to +51%; *p* = .001). As such, during the pandemic period, the monthly count of MHR ED visits averaged 177, whereas if the pandemic had not occurred, an average of 122 (Bayesian CI_95_ 114–130) would have been expected, yielding an absolute effect of +54 (Bayesian CI_95_ 47–62), as shown in eTable 4.

**Figure 3.**
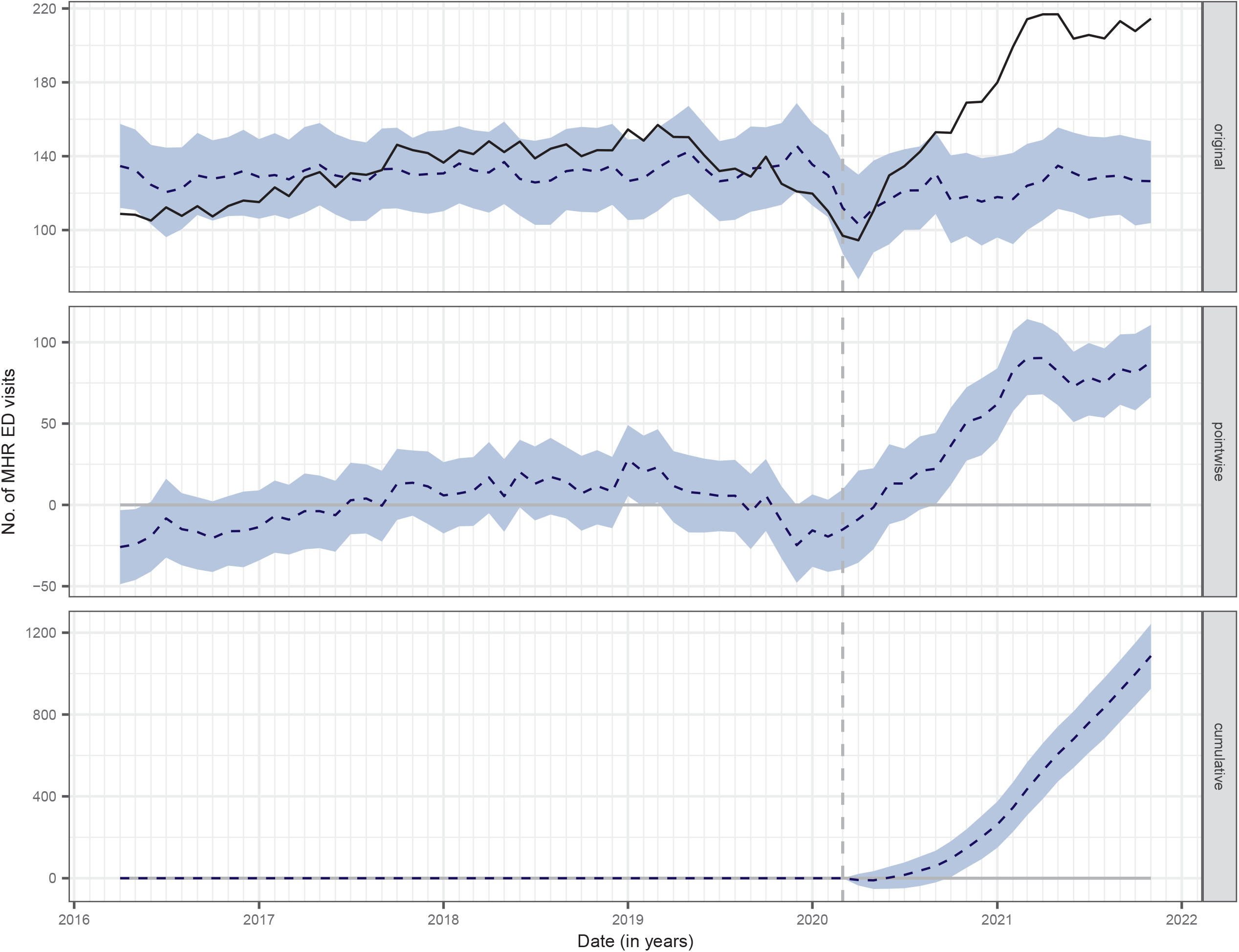
Bayesian structural time series model (using deseasonalized data) Note. Time paths of the actual and predicted values are represented by the black and blue-dotted lines, respectively. The blue shaded areas indicate 95% confidence intervals. The original panel measures the difference between actual and predicted values. The pointwise panel corresponds to the average impact of the exposure (i.e. COVID-19–associated public health measures), whereas the cumulative panel refers to the cumulative impact of the latter. Abbreviations: ED, emergency department; MHR, mental health-related.

Parametric and non-parametric GAM estimates are reported in Table 2, and represented graphically in eFigure 4. The GAM estimation indicated that the chance of presenting for a MHR vs non-MHR visit was influenced by seasonality (EDF = 7.34, p < 0.001) and age (EDF = 7.14, p < 0.001). Overall, the probability of a MHR ED visit was higher in the earlier and later months of the year (i.e. during the winter period), and tended to decrease during the summer period. However, this pattern was more pronounced during the pandemic period (EDF = 3.04, p < 0.01), as shown in eFigure 4b. The chance of presenting for a MHR ED visit increased non-linearly with age. With respect to COVID-19, the effect of age (as a continuous variable) was similar between time periods for youth up to 14 years old but differed for adolescents aged 15–17 years (EDF = 4.11, p < 0.001). More specifically, eFigure 4b shows a decrease for those aged 15–17 years during COVID-19, whereas an increase was observed among the latter before COVID-19. This result was corroborated in the sensitivity analysis treating age as a dichotomous variable (eTable 5).

**Table 2.** Semi-parametric GAM for MHR ED utilization, April 1, 2016 to November 2021.

| **Parametric terms** |  |  |  |
| --- | --- | --- | --- |
|  | **Estimate** | **SE** | **p-value** |
| **Intercept** | -3.312 | 0.039 | <0.001 |
| **Time period** |  |  |  |
| Prepandemic (ref) |  |  |  |
| Pandemic | 0.126 | 0.067 | 0.062 |
| **Sex** |  |  |  |
| Male (ref) |  |  |  |
| Female | 0.353 | 0.028 | <0.001 |
| **SES** |  |  |  |
| Most deprived | -0.267 | 0.038 | <0.001 |
| Average (ref) |  |  |  |
| Most privileged | -0.174 | 0.040 | <0.001 |
| **Sex (female): time period (pandemic)** | 0.348 | 0.048 | <0.001 |
| **SES (most deprived): time period (pandemic)** | 0.012 | 0.064 | 0.850 |
| **SES (most privileged): time period (pandemic)** | 0.089 | 0.067 | 0.184 |
| **Smooth terms** |  |  |  |
|  | **EDF** |  | **p-value** |
| **Seasonality** | 7.345 |  | <0.001 |
| **Age** | 7.137 |  | <0.001 |
| **Seasonality: time period (pandemic)** | 3.038 |  | 0.004 |
| **Age: time period (pandemic)** | 4.106 |  | <0.001 |

Note. The colon symbol is employed to represent the interaction between two terms. Age and seasonality are measured in years and months, respectively. Abbreviations: ED, emergency department; EDF, effective degrees of freedom (of the functional parameters); GAM, generalized additive model; MHR, mental health-related; SE, standard error (for the parameter estimate); SES, socioeconomic status

We present the parametric estimates as odds ratio (OR) and corresponding 95% CI in a forest plot (eFigure 4a). Female individuals were more likely to present for a MHR ED visit than male individuals, both before (OR 1.42, 95% CI 1.35–1.50) and during the COVID-19 pandemic (OR 2.02, 95% CI 1.73–2.35). By contrast, the chance was lower among youth who were the most materially deprived (OR 0.77, 95% CI 0.71–0.83 [pre-pandemic]; OR 0.78, 95% CI 0.70–0.86 [pandemic]) and those who were the most materially privileged (OR 0.84, 95% CI 0.78–0.91 [pre-pandemic]; OR 0.92, 95% CI 0.82–1.02 [pandemic]), compared to their counterparts in the average SES group.

Between the pre-pandemic and pandemic periods, the chance of presenting for a MHR ED visit increased among female individuals (OR 1.42, 95% CI 1.29–1.56). No difference by SES group during and before the COVID-19 pandemic was found (OR 1.01, 95% CI 0.89–1.15 [most deprived]; OR 1.09, 95% CI 0.96–1.25 [most privileged]).

## DISCUSSION

This retrospective cross-sectional study found that the COVID-19 pandemic period was associated with an increase in MHR ED utilization, irrespective of its overall trend and annual seasonality. Monthly counts of MHR ED visits increased by approximately 44% during COVID-19 compared to prior years. During the pandemic period, the odds of presenting for a MHR ED visit were higher among girls than those during the pre-pandemic period. Conversely, lower odds of presenting for a MHR ED visit were found in older adolescents (>15 years) during COVID-19. To our knowledge, these findings are the first to describe the magnitude of changes in MHR ED utilization in youth during COVID-19 by sociodemographic factors in Canada.

Our findings support a substantial increase in MHR ED utilization in the pandemic period compared to the pre-pandemic period, despite an initial decrease in the first three months of COVID-19. These findings corroborate with previous international studies.(7-12, 14-17, 21-24) The strongest increase appeared to take place during winter months, a period when public health measures, including school closures were stricter.(36) In Canada, and specifically in Québec, where access to primary healthcare services remains limited for many, even in non-pandemic times,(37) the loss of supportive school and community structures could have contributed to this increased utilization of ED services.

Our analyses revealed that the odds of presenting to the ED for MHR diagnoses were greater among girls, and that this was intensified by the pandemic. These results coincide with those obtained in Australia and the US.(13, 25) In fact, an increase in MHR ED utilization among adolescent girls aged 12–17 years was also seen in New South Wales during the first year of the pandemic (January 2020 to February 2021).(13) Weekly ED visits for adolescent girls also increased for eating and tic disorders in 2020-2021 and depression and obsessive-compulsive disorder in 2021 compared to 2019 in the US.(25) Conversely, while some studies have suggested an increase in MHR symptoms among girls since the onset of the pandemic,(38, 39) a repeated cross-sectional study using representative data from Ontario and Québec, Canada’s two largest provinces, showed that the increases in mental health symptoms among adolescents appeared to be similar between boys and girls. Further, these trends were no greater between the years 2018 and 2019 (pre-pandemic) than between the years 2019 and 2020 (pre/post onset of the pandemic).(40) It can thus be suggested that reasons other than changes in population-level mental health symptomatology, such as increases in help-seeking behaviours, may have contributed to the increased odds of MHR ED presentations among girls vs boys.(41)

In our study, there was a higher proportion of MHR ED visits by older (12–17 years) vs younger (5–11 years) youth, though increases in visit counts during the pandemic period could be seen in both age groups. Our findings are consistent with a recent Canadian study on healthcare services utilization for eating disorders in Ontario which showed a similar increase in MHR ED visits among younger (age 3–13 years) and older (ages 14–17 years) youths.(42) Interestingly, our non-linear estimate of the effect of age on MHR ED utilization was slightly smaller among older adolescents (ages 15–17 years) during the pandemic (vs pre-pandemic) period. Adolescence coincides with a key period of brain development and the formation of one’s personal and social identity.(43) Adolescents may be particularly vulnerable to public health preventive measures that can lead to disruptions in their social life,(44) but, as they get older, may also be able to develop new coping strategies such as connecting with peers online and outdoors, and engaging in leisure and health-promoting activities. Younger youth, who may not have the means or capacity to develop the same coping mechanisms, appear to have been equally, if not more strongly affected by school closures and other pandemic-related measures, which may have contributed to the increase in MHR ED utilization seen in this age group.(45)

Our study revealed that prior to the onset of the pandemic, MHR ED visits were less likely to be attributed to youth from both socioeconomically advantaged and disadvantaged areas when compared to those from average SES areas. Our model indicated that this same pattern remained during the pandemic. Indeed, our data showed an absolute increase in number of visits that was similar for all three SES groups, suggesting that pandemic-related effects on pediatric healthcare service utilization were similar across the SES spectrum. Our findings differ from those reported by Hu and colleagues in Australia, where increases in MHR ED utilization were higher for youth from socioeconomically advantaged areas.(13) Though increased utilization of MHR ED services may serve as an indicator of worsening youth mental health in some circumstances, this indicator alone does not adequately capture the exacerbation of existing healthcare inequalities among youth from lower vs higher SES, and may be more reflective of SES-related differences in access to appropriate services.(46, 47)

## Limitations

There are limitations to our study. First, our findings are based on a single, large, tertiary pediatric center, and might not be generalizable to other clinical settings. However, this center is the largest pediatric hospital with the greatest number of ED visits in the province. Second, the comprehensiveness of the outcome and patient characteristics was limited by data availability. We could only identify the primary diagnosis as recorded by the treating physician at the ED (without confirmation by a mental health provider), despite it being common for patients to present multiple diagnoses. Third, when considering SES, the geocoding of postal codes could not provide patient-level data and resulted in missing data (∼5% overall). Use of census information on dissemination areas from 2016, rather than the exact year during which the ED visits occurred, might have introduced bias. Finally, MHR ED utilization (as measured by the monthly number of visits) offers only a single viewpoint on youth mental health, and should therefore be considered in the larger context of other physician- and non-physician-based mental healthcare services utilization.(2) Increased MHR ED utilization could also be a reflection of other societal disruptions, such as the loss of access to other school- and community–based services.(48)

## CONCLUSIONS

The COVID-19 pandemic has placed an unprecedented stress on pediatric mental health services as shown by abrupt increases in MHR ED visits. While EDs can serve an important purpose in providing rapid mental health services for youth in situations of crisis, there is a need for sustained, targeted and scalable mental health resources to support youth mental health. This appears to be especially true for girls and should be considered in preparation for future public health crises.

## Data Availability

The datasets used and/or analysed during the current study are available from the corresponding author on reasonable request.

## List of abbreviations

CHU: centre hospitalier universitaire
COVID-19: coronavirus disease 2019
ED: emergency department
GAM: generalized additive model
GLM: generalized linear model
ITS: interrupted time series
MHR: mental health–related
REML: restricted maximum likelihood
SES: socioeconomic status
STROBE: Strengthening the Reporting of Observational Studies in Epidemiology
UK: United Kingdom
US: United States

## DECLARATIONS

### Ethics approval and consent to participate

Ethical approval was obtained from the CHU Sainte-Justine Research Ethics Committee (Protocol ID: MP-21-2021-2930).

### Consent for publication

All patient data were deidentified; thus, informed consent was not required.

### Authors’ contributions

NC and OD conceived the study. GB and NC designed the study. JG oversaw the data collection and AS performed the data extraction. OD obtained ethical approval. GB had full access to all data and conducted the analyses, with feedback from AB. GB and NC drafted the manuscript. AB, MO and MCG revised the manuscript for important intellectual content. All authors contributed to the interpretation of data and read and approved the final manuscript.

## Acknowledgements

Not applicable.

