## Supplemental material for "A comparative analysis of pediatric mental health-related emergency department utilization in Montréal, Canada before and during the COVID-19 pandemic"

**Supplementary material**

**eTable 1** MHR ED diagnoses included in the study sample
**eTable 2.** Patient characteristics of included vs. excluded MHR ED visits
**eTable 3.** Frequency of the number of MHR ED visits per unique patient
**eTable 4** Posterior estimates of the causal impact of COVID-19 on MHR ED visits, April 1, 2016 to November 30, 2021
**eTable 5.** Semi-parametric GAM for MHR ED utilization, April 1, 2016 to November 30, 2021 (with age group as a dichotomous variable)
**eFigure 1.** Bayesian structural time series model (with raw data)
**eFigure 2.** Decomposition of additive time series
**eFigure 3.** Original and deseasonalized time series
**eFigure 4.** Graphical representation of the parametric and smooth terms included in the semi-parametric GAM

**eTable 1. MHR ED diagnoses included in the study sample**

| Diagnostic categories | MHR ED diagnoses |
|---|---|
| Abuse/neglect | Possibilité abus sexuel;<br>Possibilité de viol – possibilité agression sexuelle;<br>Possibilité enfant maltraité, possibilité abus physique;<br>Trouble social, psychosocial, trouble psych. |
| Anxiety disorders | Angoisse / crise d'angoisse, attaque de panique;<br>Anxiété, trouble anxieux, stress;<br>Factice, trouble factice, simulation, conversion;<br>Phobie;<br>Phobie scolaire;<br>Somatisation;<br>Trouble de l'adaptation, problème d'adaptation;<br>Trouble psychosomatique, problème psychosomatique. |
| Eating disorders | Anorexie mentale;<br>Anorexie, anorexie nerveuse;<br>Trouble alimentaire, problème alimentaire. |
| Mood disorders | Dépression, trouble dépressif;<br>Humeur triste;<br>Maladie bipolaire, trouble bipolaire, maniaco-dépression;<br>Trouble de l'humeur, problème d'humeur. |
| Other mental health | 19 – Psychiatrie;<br>Agitation;<br>Agressivité;<br>Irritabilité;<br>Psychose;<br>Psychose maniaco-dépressive, PMD;<br>Syndrome Gilles de la Tourette, Gilles de la Tourette;<br>Terreur nocturne;<br>Tics;<br>Trouble du comportement, crise, problème de comportement;<br>Trouble du sommeil;<br>Troubles mentaux, suivi en pédo-psy. |
| Substance use | 20 – Dépendance;<br>Intoxication alcool;<br>Intoxication amphétamines;<br>Intoxication champignons;<br>Intoxication cocaïne;<br>Intoxication drogues de rue;<br>Intoxication THC;<br>Intoxication tricycliques. |
| Suicidal ideation | Automutilation;<br>Idées suicidaires, suicidaires;<br>Intoxication acétaminophène;<br>Intoxication aspirine;<br>Intoxication AAS;<br>Intoxication méthanol;<br>Intoxication, intox;<br>Tentative suicidaire. |

Note. MHR ED diagnoses are listed in French, as recorded in the medical system by the treating physicians.
Abbreviations. ED, emergency department; MHR, mental health-related.

**eTable 2. Patient characteristics of included vs. excluded MHR ED visits**

| | Included (N = 9308) | Excluded (N = 440) | p-value |
|---|---|---|---|
| **Age** | | | |
| Median (IQR) | 14.3 (11.4–16.0) | 14.7 (12.4–16.4) | 0.018 |
| **Age group** | | | |
| 5–11 years | 2661 (28.6%) | 2763 (28.3%) | 0.049 |
| 12+ years | 6647 (71.4%) | 6986 (71.7%) | |
| **Sex** | | | |
| Female | 5816 (62.5%) | 6106 (62.6%) | 0.349 |
| Male | 3492 (37.5%) | 3642 (37.4%) | |

Note. Between-group differences were assessed using Kruskal-Wallis (for continuous variables) and $\chi^2$ tests (for categorical variables).
Abbreviations. ED, emergency department; MHR, mental health-related.

**eTable 3. Frequency of the number of MHR ED visits per unique patient**

| Number of MHR ED visits | Number of unique patients | Relative frequency (%) |
|---|---|---|
| 1 | 6369 | 82.9 |
| 2 | 942 | 12.3 |
| 3 | 222 | 2.9 |
| 4 | 78 | 1.1 |
| 5 | 36 | 0.5 |
| 6 | 19 | 0.3 |
| 7+ | 20 | 0.3 |

Note. Some categories are pooled to ensure patient confidentiality.
Abbreviations. ED, emergency department; MHR, mental health-related.

**eTable 4. Posterior estimates of the causal impact of COVID-19 on MHR ED visits, April 1, 2016 to November 30, 2021**

| | Time series data | | | |
|---|---|---|---|---|
| | Raw | | Deseasonalized | |
| | Average | Cumulative | Average | Cumulative |
| **Actual** | 172 | 3437 | 177 | 3531 |
| **Prediction** (95% CI) | 101 (86–118) | 2028 (1721–2354) | 122 (114–130) | 2445 (2285–2597) |
| **Absolute effect** (95% CI) | 70 (54–86) | 1409 (1083–1716) | 54 (47–62) | 1086 (934–1246) |
| **Relative effect** (95% CI), % | 69 (53–85) | 69 (53–85) | 44 (38–51) | 44 (38–51) |

Note. Numbers have been rounded off to whole numbers, as have percentages.
Abbreviations: ED, emergency department; MHR, mental health-related.

**eTable 5. Semi-parametric GAM for MHR ED utilization, April 1, 2016 to November 30, 2021 (with age group as a dichotomous variable)**

| | Parametric terms | | |
|---|---|---|---|
| | **Estimate** | **SE** | **p-value** |
| **Intercept** | -3.784 | 0.041 | <0.001 |
| **Time period** | | | |
| Prepandemic (ref) | | | |
| Pandemic | 0.168 | 0.072 | 0.019 |
| **Age group** | | | |
| 5–11 years (ref) | | | |
| 12+ years | 1.633 | 0.029 | <0.001 |
| **Sex** | | | |
| Male (ref) | | | |
| Female | 0.393 | 0.028 | <0.001 |
| **SES** | | | |
| Most deprived | -0.281 | 0.038 | <0.001 |
| Average (ref) | | | |
| Most privileged | -0.167 | 0.040 | <0.001 |
| **Age group** (12+ years): **time period** (pandemic) | -0.041 | 0.051 | 0.423 |
| **Sex** (female): **time period** (pandemic) | 0.334 | 0.048 | <0.001 |
| **SES** (most deprived): **time period** (pandemic) | 0.015 | 0.064 | 0.814 |
| **SES** (most privileged): **time period** (pandemic) | 0.082 | 0.067 | 0.223 |
| | **Smooth terms** | | |
| | **EDF** | | **p-value** |
| **Seasonality** | 7.353 | | <0.001 |
| **Seasonality**: **time period** (pandemic) | 3.136 | | 0.002 |

Note. The colon symbol is employed to represent the interaction between two terms. Seasonality is measured in months.
Abbreviations: ED, emergency department; EDF, effective degrees of freedom (of the functional parameters); GAM, generalized additive model; MHR, mental health-related; SE, standard error (for the parameter estimate); SES, socioeconomic status.

**eFigure 1. Bayesian structural time series model (with raw data)**

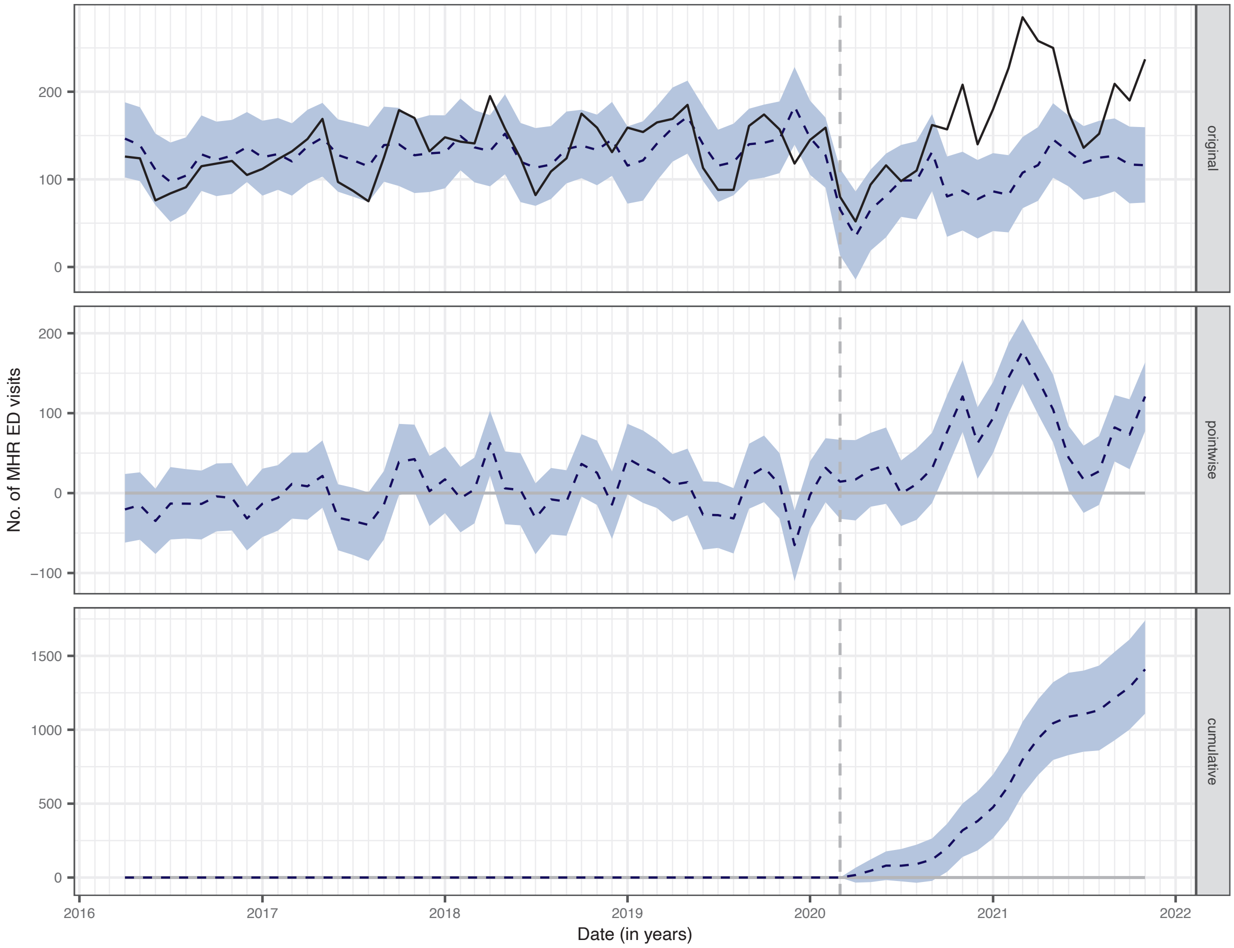


Note. Time paths of the actual and predicted values are represented by the black and blue-dotted lines, respectively. The blue shaded areas indicate 95% confidence intervals. The original panel measures the difference between actual and predicted values. The pointwise panel corresponds to the average impact of the exposure (i.e. COVID-19–associated public health measures), whereas the cumulative panel refers to the cumulative impact of the latter.

**eFigure 2. Decomposition of additive time series**

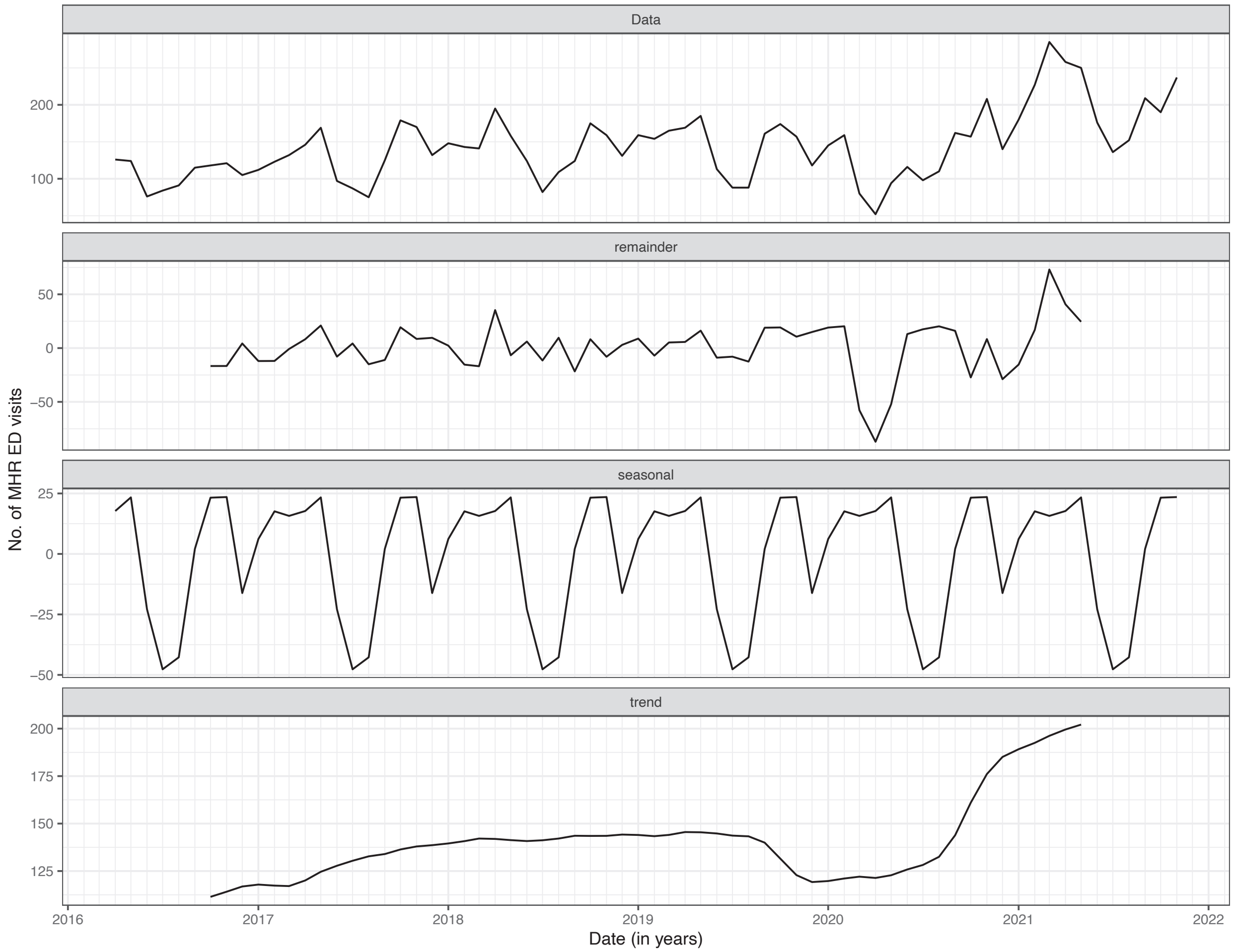


Note. The data panel represents the raw data on the number of ED MHR visits between April 2016 and November 2021. The remainder panel highlights atypical events in the time series (i.e. outliers that cannot be explained alone by the long-term trend and seasonality). The seasonal panel measures the time series' seasonality, whereas the trend panel indicates the long-term trend of ED MHR visits.

**eFigure 3. Original and deseasonalized time series**

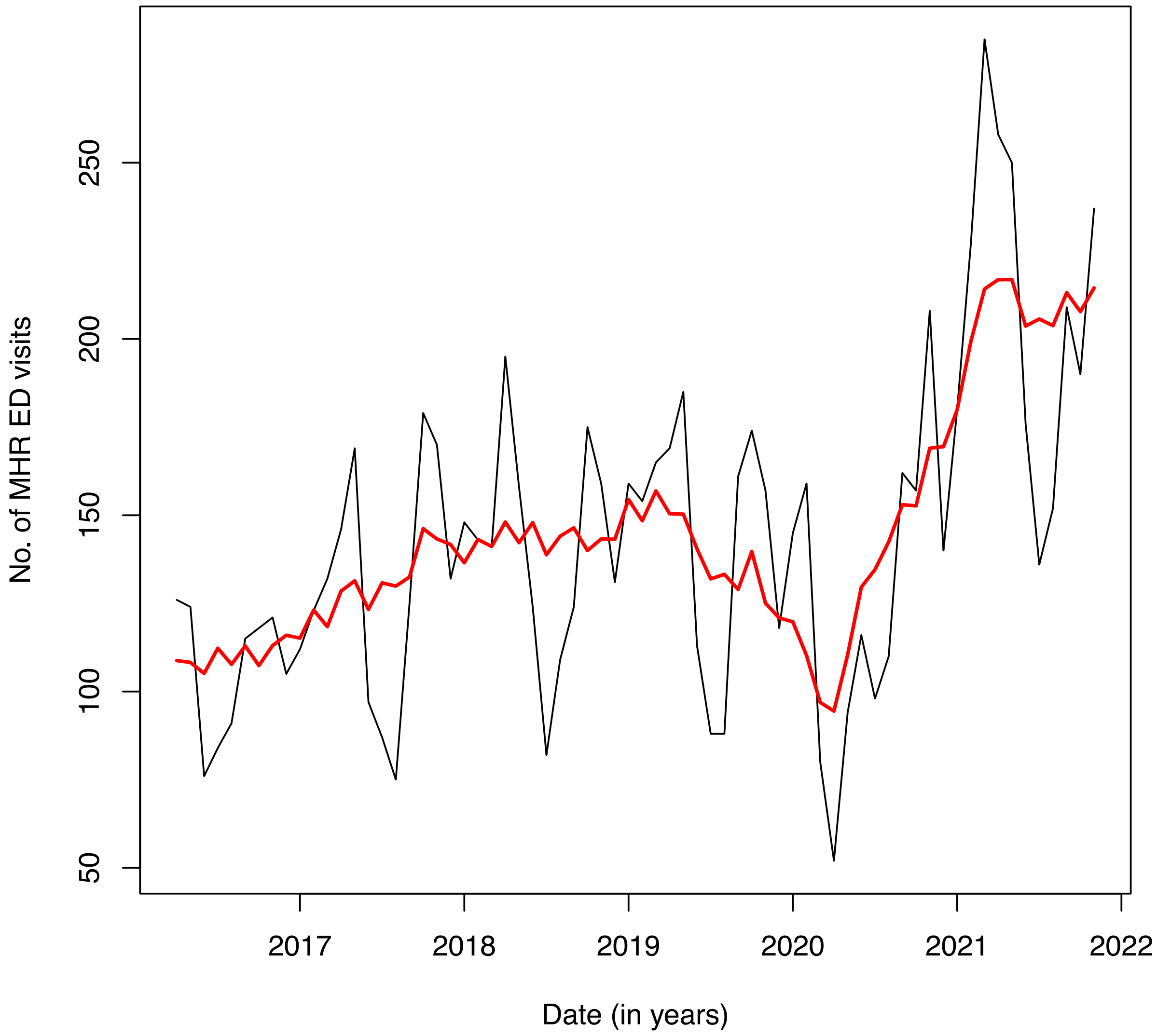


Note. The black line represents the raw data (original time series), whilst the red line estimates the adjusted data (deseasonalized time series).

**eFigure 4. Graphical representation of the parametric and smooth terms included in the semi-parametric GAM**

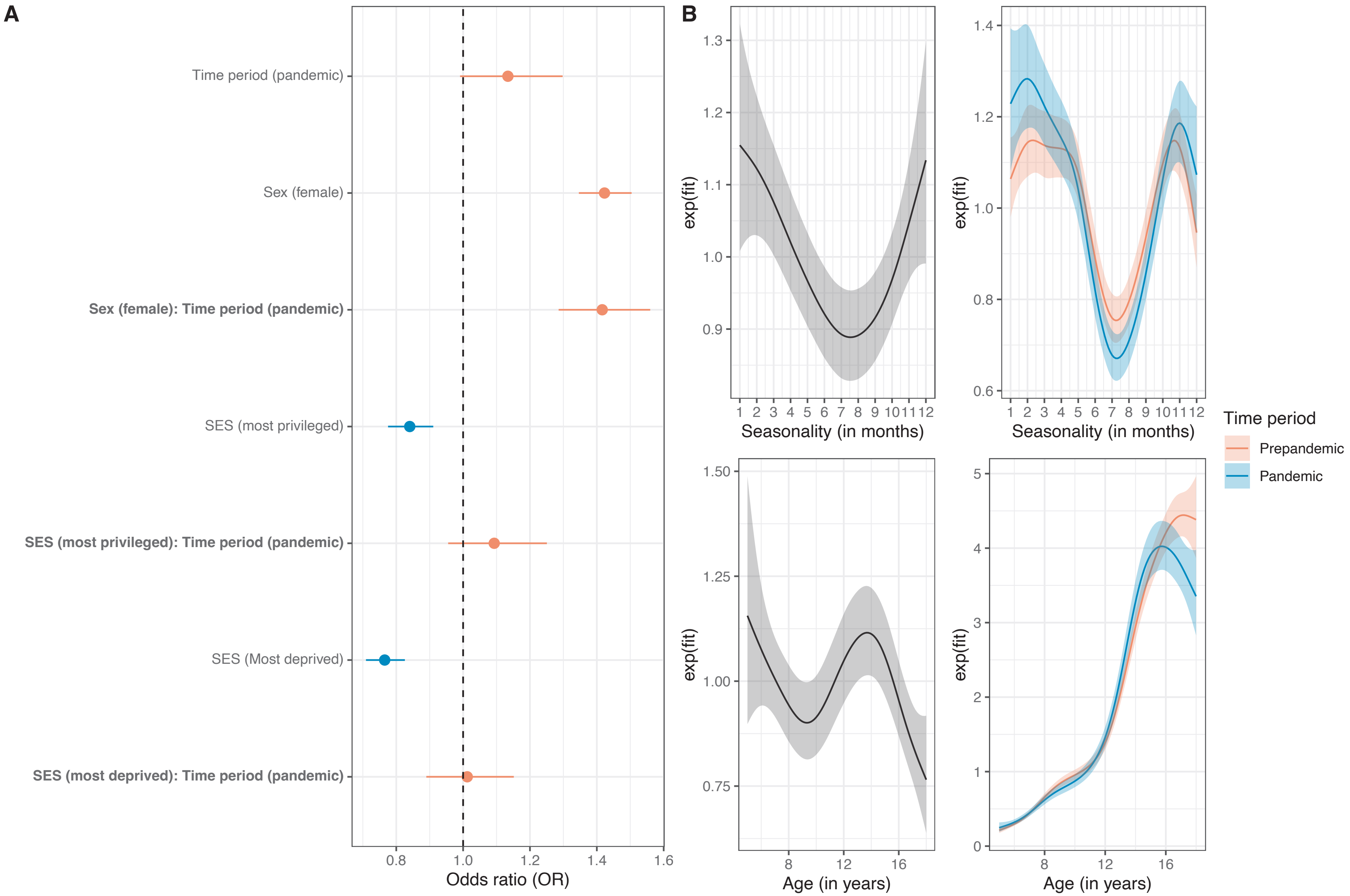


Note. Panel A is a forest plot of the parametric terms included in the GAM. Panel B shows the effect of seasonality and age, respectively, by time period on MHR ED utilization. Shaded areas indicate 95% CIs.
Abbreviation: GAM, generalized additive model.
